# The OncoSim-Breast cancer microsimulation model

**DOI:** 10.1101/2020.05.22.20110569

**Authors:** J.H.E. Yong, C. Nadeau, W. Flanagan, A. Coldman, K. Asakawa, R. Garner, N. Fitzgerald, M. Yaffe, A.B. Miller, on behalf of the OncoSim-Breast Working Group

**Author notes:** Corresponding author: Jean Hai Ein Yong, Canadian Partnership Against Cancer, 145 King St. W, Toronto, M5H 1J8 Ontario, Canada.

## Abstract

**Background:** The increasing demand for health care resources requires measures to evaluate the impact of cancer control approaches. A cancer simulation model can help integrate new knowledge to inform clinical and policy decisions. OncoSim-Breast is a breast cancer simulation model. This paper aims to describe the key assumptions in the OncoSim-Breast model and how well it reproduces more recent breast cancer trends and the observed effects in a randomized screening trial.

**Methods:** The OncoSim-Breast model simulates the onset, growth and spread of invasive and ductal carcinoma in situ tumours. The model combines Canadian cancer incidence, mortality, screening program and cost data to project population-level outcomes. Users can change the model input to answer specific policy questions. Here we report three validation exercises. First, we compared the model’s projected breast cancer incidence and stage distributions with the observed data in the Canadian Cancer Registry. Second, we compared OncoSim’s projected breast cancer mortality with the Vital Statistics. Third, we replicated the UK Age trial to compare the model’s projections with the trial’s observed screening effects.

**Results:** OncoSim-Breast’s projected incidence, mortality and stage distribution of breast cancer were close to the observed data in the Canadian Cancer Registry and the Vital Statistics. OncoSim-Breast also reproduced the breast cancer screening effects observed in the UK Age trial.

**Interpretation:** OncoSim-Breast’s ability to reproduce the observed population-level breast cancer trends and the screening effects in a randomized trial increases the confidence of using its results to inform policy decisions related to early detection of breast cancer.

## INTRODUCTION

Rapidly emerging knowledge in breast cancer control has put pressure on the health system for the adoption of new technologies and policies. Randomized trials are the gold standard of evidence to introduce new interventions in clinical practice and public health; however, such evidence is not always relevant for informing policy decisions because the context of the interventions evolves quickly compared to the time that elapses between the design of a trial and the availability of its results. For example, most breast cancer screening randomized trials were from the era before breast cancer adjuvant treatment was available and used film-screen mammography^1,2^; breast cancer survival has since vastly improved^3^ and digital mammography has superseded film-screen mammography. A cancer simulation model can help integrate evidence from multiple sources and make them more relevant to inform contemporary clinical and policy decisions. Several groups have developed sophisticated cancer-specific models based on the natural history of cancer that can be revised for additional analyses and incorporate knowledge from experts in different areas^4^.

OncoSim is an example of a cancer simulation model. The validation and applications of OncoSim colorectal, cervical, and lung cancers have been described previously^5-19^. Breast cancer is the latest addition to OncoSim’s suite of cancer models. The primary objective of OncoSim-Breast is to investigate emerging issues related to breast cancer control in Canada. This work builds on a strong foundation of analyses performed over a decade ago to estimate the impact of diagnostic and therapeutic approaches to non-metastatic breast cancer in Canada, using the Statistics Canada POHEM mathematical model^20^. The present paper has two goals. First, it aims to describe the key assumptions in OncoSim-Breast. Secondly, it compares OncoSim-Breast’s projections with observed data in the Canadian Cancer Registry, projected breast cancer mortality estimates in the Canadian Vital Statistics, and the observed screening effects in a randomized trial.

## METHODS

### Overview of OncoSim web application

With funding from Health Canada, OncoSim is led and supported by the Canadian Partnership Against Cancer, with model development by Statistics Canada, to help answer complex questions related to Canada’s cancer control. OncoSim was developed by experts in statistics, microsimulation, cancer screening, clinical epidemiology, oncology, and health economics. It is unique in that it is available at no charge to users in the public sector. The model comes with pre-populated inputs to facilitate analyses. Through an online platform with 24/7 access, users can create unique scenarios by modifying one or more of the model inputs and export OncoSim’s projections for further analyses. In addition to peer-reviewed publications, its projections have been used to inform clinical practice guidelines, Canadian Cancer Statistics reports, and other national and jurisdictional-level cancer control decisions^21-25^. An earlier paper describes details about OncoSim’s development, validation, calibration approach and applications.^13^

### OncoSim-Breast

The model inputs have several components:

- Demography
- Natural history
- Cancer detection
- Screening
- Disease progression
- Breast cancer costs
- Impact of breast cancer on health-related quality of life

The model combines these inputs to project outcomes, such as breast cancer incidence and mortality, screening outcomes, stage and age at diagnosis, life-years, quality-adjusted life-years, lifetime breast cancer costs, and screening or follow-up procedure costs (Figure 1).

**Figure 1.**
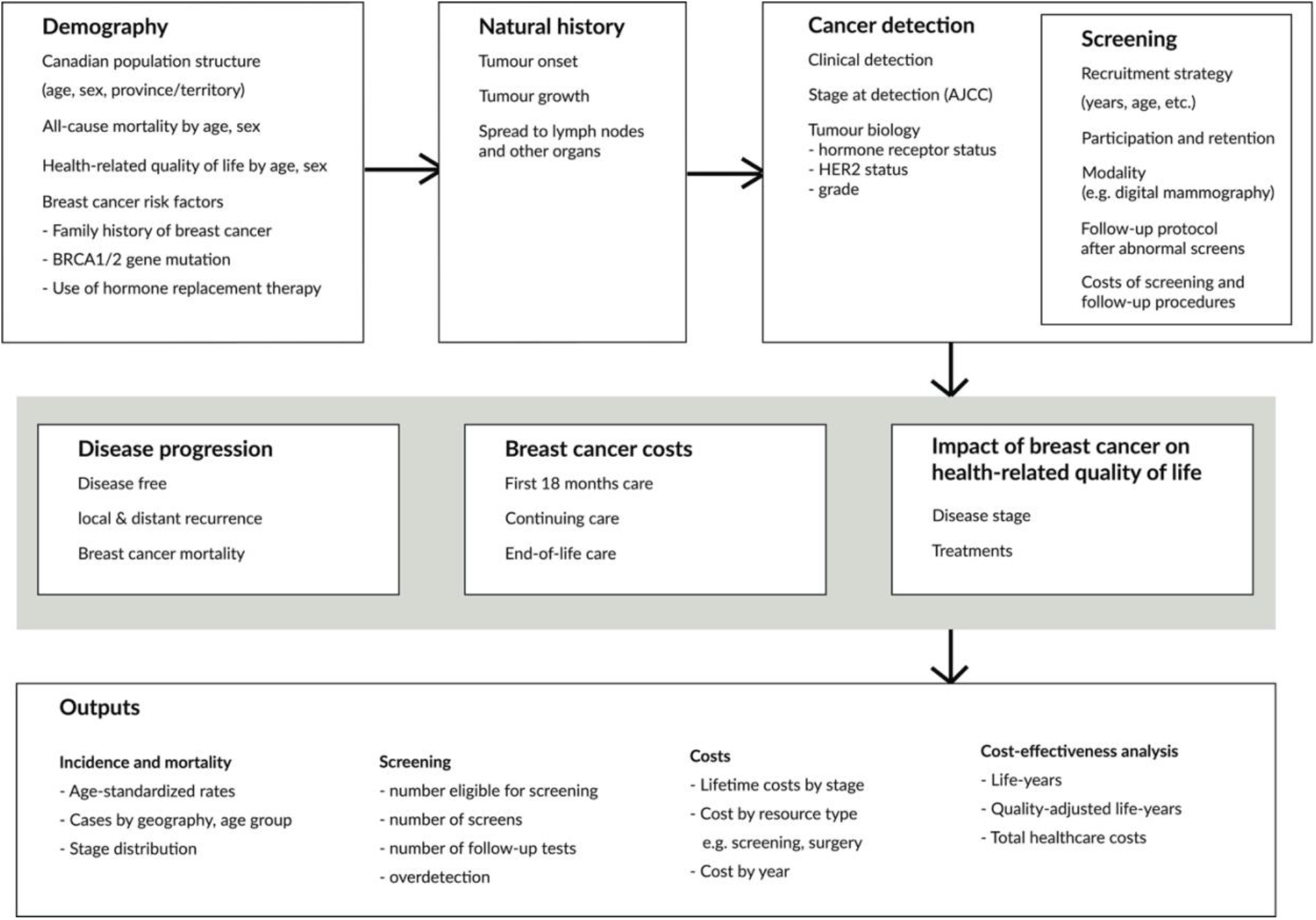
Schematic diagram of the OncoSim-Breast model.

#### Demography

OncoSim simulates one individual at a time, replicating the age and sex distributions, and all-cause mortality of the population in each province and territory in Canada (Supplemental Appendix 1). Each simulated individual has attributes, such as demography (sex, province/territory), and breast cancer-related risk factors (BRCA1/2 gene mutation, family history, exposure to hormone replacement therapy, Supplemental Table 1).

#### Natural history

OncoSim-Breast simulates the onset, growth and spread of tumours, both invasive cancer and Ductal Carcinoma in situ (DCIS) (Supplemental Appendix 2). Invasive tumours can develop without a prior DCIS; DCIS and invasive tumours can also develop and grow independently of each other. Thus, a woman could have one of the four outcomes: (i) a DCIS tumour, (ii) an invasive tumour, (iii) a DCIS tumour that becomes invasive and evolves independently of the initial DCIS, or (iv) no breast tumour at all. The development, tumour biology, growth, and clinical detection of breast cancers, both invasive cancer and DCIS, were calibrated from inputs in the University of Wisconsin Breast Cancer Epidemiology Simulation Model (“Wisconsin Breast model”)^26^ to match the incidence of cancer by age group and year in the National Cancer Incidence Reporting System (1969-1991), Canadian Cancer Registry (1992-2013) and Canadian Breast Cancer Screening Database (2007-2008).

**Tumour onset:** In OncoSim-Breast, tumours start from 2mm, based on the probable minimum size detectable by mammography screening and similar to the Wisconsin Breast model. Probability of tumour onset varies by age and years (Supplemental Figure 1). In addition, the risk increases if a woman has any of the breast cancer-related risk factors (BRCA1/2 mutation, family history of breast cancer or exposure to hormone replacement therapy, Supplemental Tables 3-4); if a woman has previously had a DCIS tumour, she is also more likely to have an invasive cancer.

**Tumour growth:** In the model, tumours grow according to the time since tumour onset. the presence of BRCA1/2 gene mutation, tumour type (DCIS or invasive) and tumour aggressiveness (Supplemental Figure 2).

**Tumour spread:** An invasive tumour can spread into lymph nodes and beyond the breast. The spread to other lymph nodes is determined by the size and growth rate of the tumour, and time since tumour onset. The tumour size and the number of lymph nodes affected then determine if invasive cancer spreads beyond the breast (metastasis).

#### Cancer detection, staging and tumour biology

Larger tumours are more likely to get detected clinically than smaller tumours (Supplementary Table 7). The stage at detection uses the American Joint Committee on Cancer (AJCC)’s classification of tumour size (T), nodal status (N) and metastasis (M). The tumour size and nodal status at detection are estimated using the tumour size and number of positive nodes generated from the natural history component and age. Similar to other breast cancer simulation models3, OncoSim-Breast assigns tumour biology (hormone receptor status, HER2neu status, and grade) to the tumour based on tumour size, nodal involvement, metastatic status, the age at which it is detected, and BRCA1/2 gene mutation (Supplemental Appendix 3).

#### Screening

For evaluating screening strategies or related performance, the model allows users to create different screening strategies and scenarios by modifying the following input parameters (Supplemental Appendix 4):

- Screening program recruitment strategy (e.g. start/end age and years)
- Screening participation and retention
- Screening frequency
- Screening modality (e.g. digital mammography)
- Sensitivity and specificity of screening
- Follow-up protocol after abnormal screening results
- Costs of screening and follow-up procedures

The model also includes historical breast screening trends in Canada (starting in 1986) to match the observed screening patterns reported in the screening programs in 2007-2012. Screening interventions can vary by family history and BRCA1/2 gene mutation. The model includes different screening modalities and allows their performance to vary by tumour size, age group and screen sequence (Supplemental Figure 5 and Supplemental Table 8). Women with an abnormal mammogram receive additional workups, such as diagnostic imaging, biopsy and fine-needle aspiration. The model includes costs of screening and follow-up procedures from the perspective of a public healthcare payer, such as the Ministry of Health (Supplemental Table 9). Oncosim-Breast captures the benefits of screening on breast cancer survival using lead time calibrated from observed survival data (Supplemental Figure 3).

#### Disease progression

Upon cancer detection, the model estimates the survival outcomes based on stage, tumour biology, age at diagnosis, and detection method; in the case of disease progression, it draws time to recurrence and possible breast cancer death (Supplemental Appendix 5). Stage-specific recurrence risks and breast cancer survival outcomes were estimated using data from the British Columbia Cancer Agency because comprehensive staging data only became available recently in the Canadian Cancer Registry. To capture provincial differences in stage-specific survival, the model applies province-specific relative risks, estimated from more recent data in the Canadian Cancer Registry, to the British Columbia survival curves.

#### Breast cancer costs

The model included healthcare costs associated with breast cancer from the perspective of a public healthcare payer, such as the Ministry of Health (Supplemental Appendix 6). The costs included breast cancer surgery, radiation treatment, chemotherapy, imaging tests and oncology physician fees, acute hospitalizations, emergency department visits, home care, long-term care, complex continuing care, and others. The model captures lifetime costs of breast cancer across three phases of care (first 18 months after diagnosis, continuing care and terminal care), a similar approach used in other established breast cancer simulation models.^3^

#### Health-related quality of life

To calculate quality-adjusted life-years, the model multiplies the duration of each health state with age and sex-specific preference scores for the Canadian population^27^ and breast cancer-specific health state utilities^28^ (upon cancer diagnosis) (Supplemental Appendix 7). When an individual is in several health states at the same time, we assumed the utility score is multiplicative.^29^

### Model validation

We validated our model in three ways. First, we compared the projected incidence and stage distribution of breast cancer in Canada with the observed data in the Canadian Cancer Registry (1992-2017). Second, we compared OncoSim’s projected breast cancer deaths in 2018 with the latest breast cancer death data in the Canadian Vital Statistics^30^. Lastly, as an external validation exercise, we replicated the screening strategies of the UK Age trial^31^ in OncoSim to compare OncoSim’s projected impact of breast cancer screening on incidence and mortality with the observed effects in the trial.

The UK Age trial is a randomized trial that compared annual screening in women aged 40-49 years with usual care in the UK in the 1990S^31^. Other established breast cancer simulation models have validated their predictions against the UK trial results^34^. We simulated a cohort of women born in 1950-1957 to match the birth cohort in the UK Age trial in two scenarios: (i) no screening; and (ii) annual screening for women age 40-49. In the screening scenario, we calibrated the rescreening rate to the average number of mammograms per woman in the Age trial (4.8)^33^. For each scenario, we estimated the incidence of breast cancer and breast cancer deaths in women aged 40-49 years. We then compared OncoSim Breast’s projected incidence of breast cancer (DCIS and invasive cancers) with the trial’s mean estimate and its 95% confidence interval. For breast cancer mortality, we compared the mortality reduction ratio from OncoSim Breast with the trial’s mean estimate and 95% confidence interval over a 17 years follow-up period. We chose to compare rate ratios rather than rates because the populations were different: volunteers in the UK Age trial vs. Canadian population.

## RESULTS

OncoSim’s projected incidence, stage distribution and mortality of breast cancer were close to the observed data in the Canadian Cancer Registry and the Vital Statistics^32^. Figures 2A and 2B compare OncoSim’s projected incidence rates of breast cancer (invasive cancer and DCIS) by age group in 1992-2013 with the observed data in the Canadian Cancer Registry. Figure 3 compares OncoSim’s projected incidence of breast cancer by province in 2008-2017 with the observed data in the Canadian Cancer Registry. Figure 4 compares OncoSim’s projected stage distribution in 2011-2015 with the observed data in the Canadian Cancer Registry. Lastly, OncoSim’s projected breast cancer deaths in 2018 was also close to the observed data in the Vital Statistics (27 per 100,000 women in OncoSim version 3.3.6 vs. 28 per 100,000 women in the Vital Statistics)^30^.

**Figure 2A.**
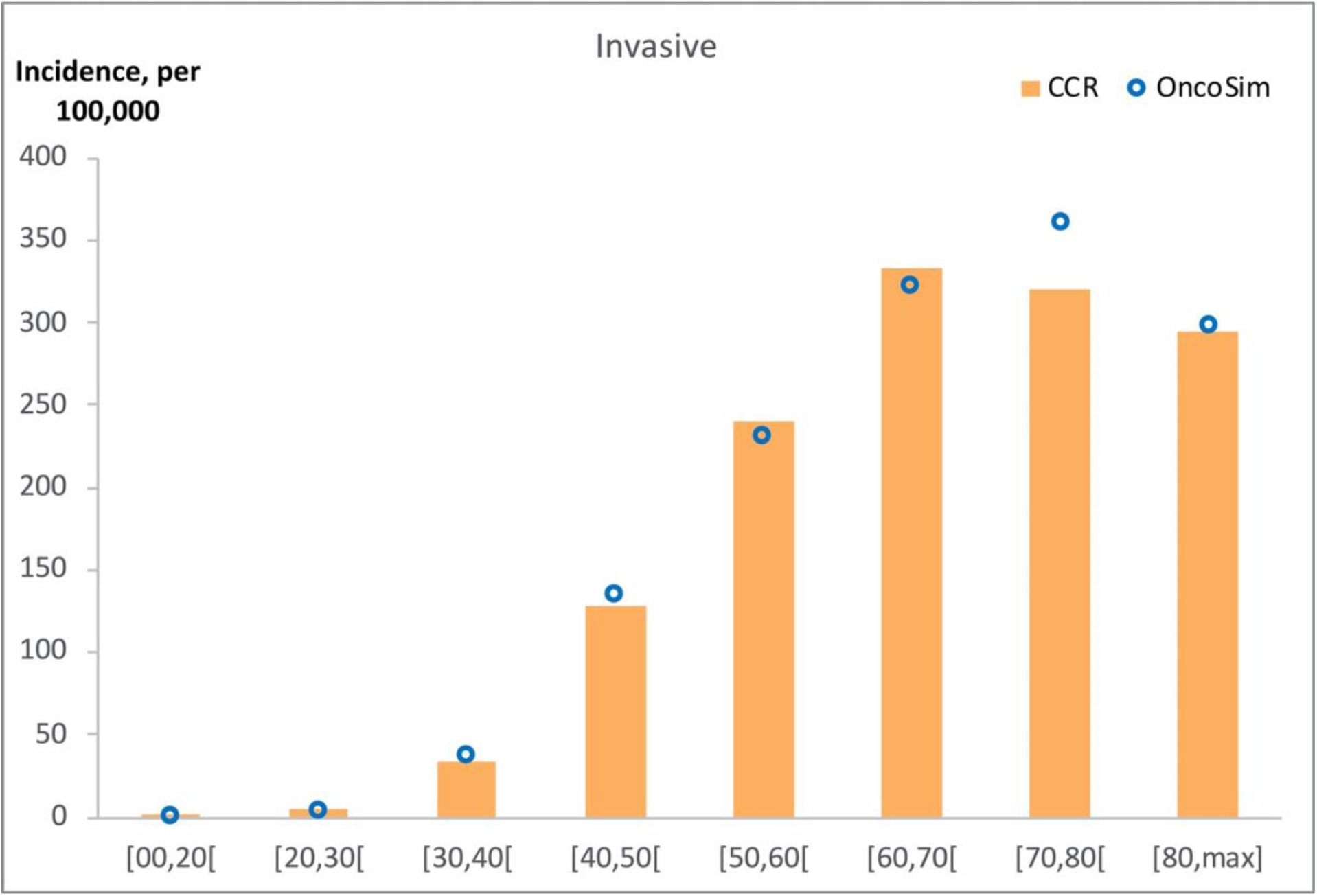
Incidence of invasive breast cancer (per 100,000 women) by age group in 1992-2013, OncoSim-Breast vs. Canadian Cancer Registry (CCR)

**Figure 2B.**
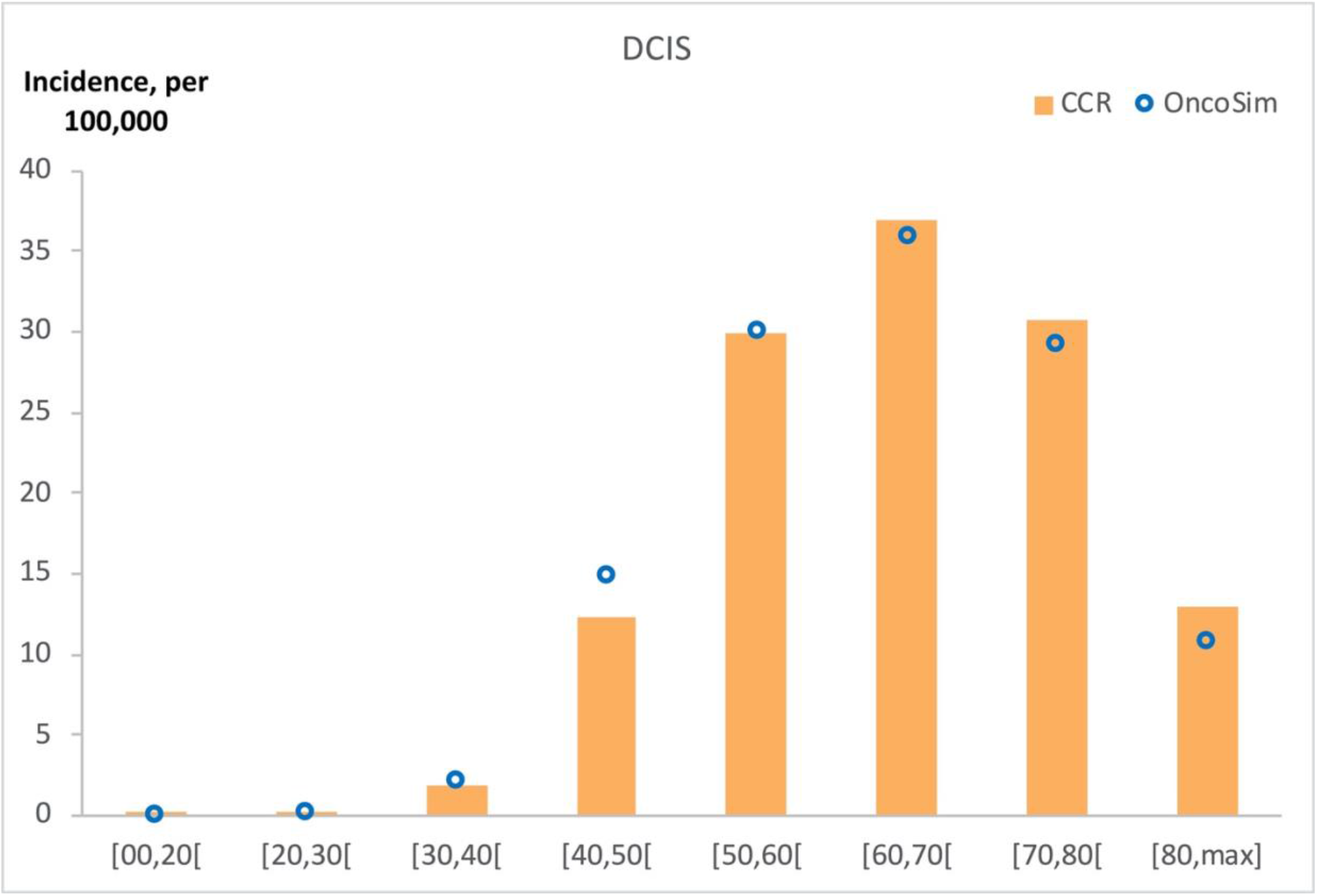
Incidence of ductal carcinoma in situ (per 100,000 women) by age group in 1992-2013, OncoSim-Breast vs. Canadian Cancer Registry (CCR)

**Figure 3.**
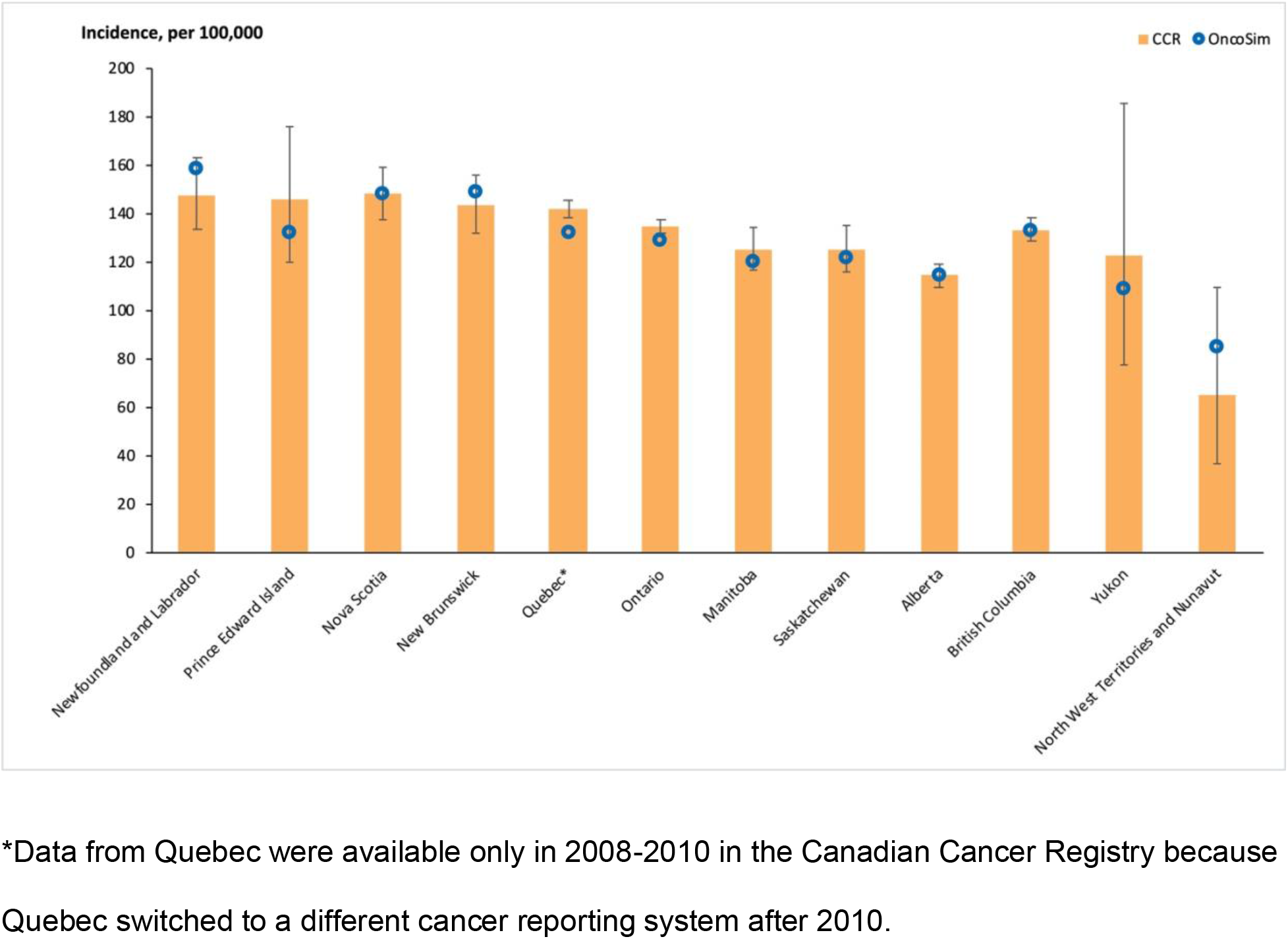
Incidence of invasive breast cancer (per 100,000 women), average per year (2008-2017), by province, OncoSim-Breast vs. Canadian Cancer Registry (CCR)

**Figure 4.**
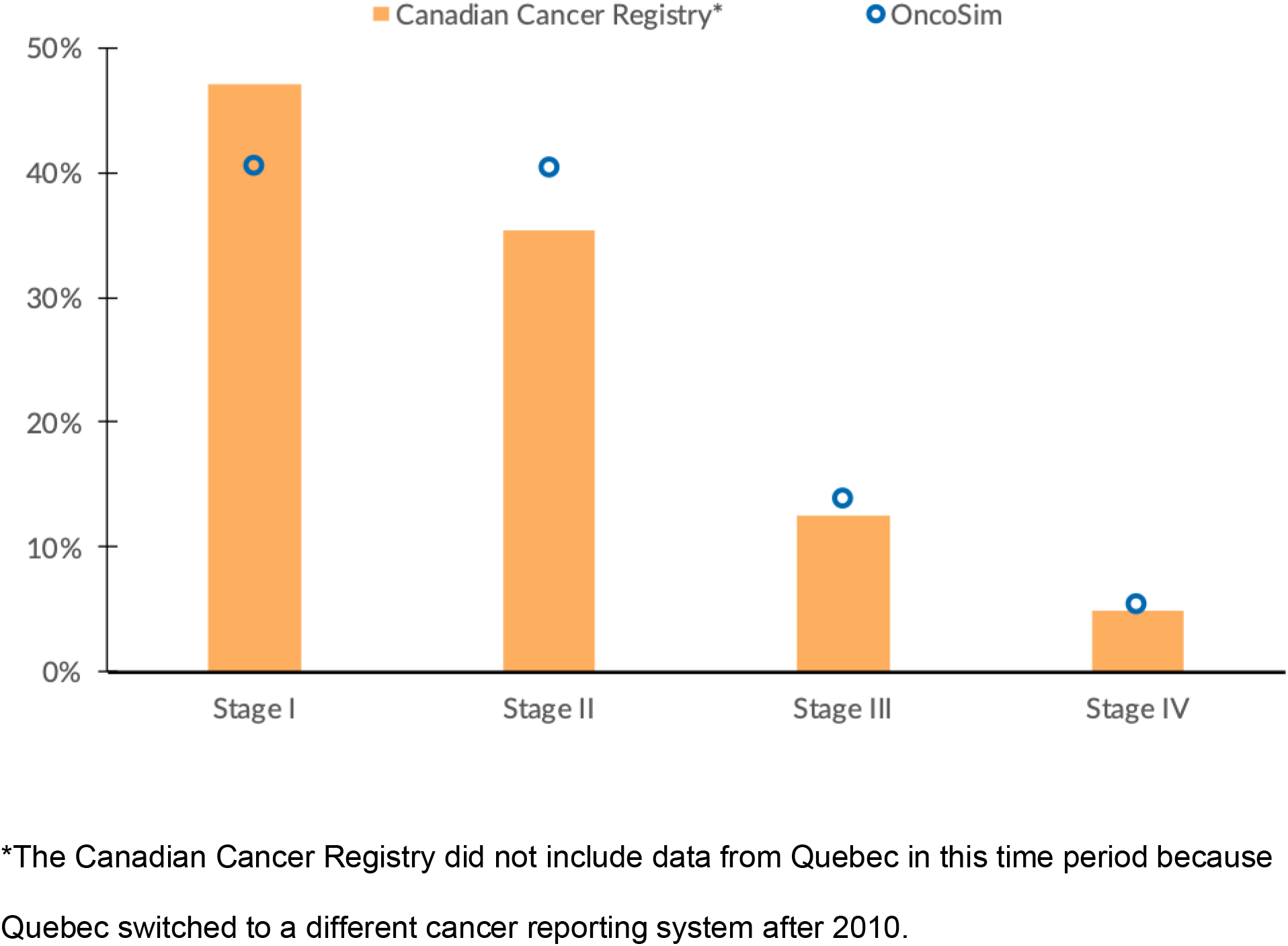
Distribution of breast cancer by stage at diagnosis, females, Canada, 2011-2015, OncoSim-Breast vs. Canadian Cancer Registry

In our external validation exercise simulating the UK Age trial, OncoSim projected that annual breast cancer screening in women age 40-49 years would detect 16% more invasive breast cancers vs. 10% (95% CI: 0.95-1.21) observed in the UK Age trial^34^. When estimating the impact of screening on breast cancer deaths, OncoSim’s projection was also within the confidence interval of the observed trial estimate. Over a 17 years follow-up period, OncoSim estimated that annual breast cancer screening in women age 40-49 years had an 85% relative risk of breast cancer death (i.e. 15% reduction), as compared with 88% (95% CI: 74%-104%) reported in the UK Age trial.^32-33^

## INTERPRETATION

This paper provides an overview of OncoSim-Breast inputs, assumptions, breast cancer cost projections and model validation results. When projecting incidence, mortality and stage at diagnosis of breast cancer, OncoSim-Breast’s estimates were close to the estimates reported in the Canadian Cancer Registry and Vital Statistics. In addition, OncoSim-Breast’s ability to reproduce the observed effects of annual breast cancer screening in a randomized screening trial increases the confidence of using the model results to inform breast cancer screening-related policy decisions.

Building upon the experience of other OncoSim models and another established breast cancer microsimulation model^3^, OncoSim-Breast was developed using Canadian data. While the model has many potential applications, its primary purpose was to evaluate the impact of interventions related to early detection, such as promoting breast cancer awareness through professional and public education and screening. For screening, the model has many detailed outputs for informing policy decisions, including the harm of screening (e.g. false-positives and overdetection), healthcare costs, and benefits (life-year gained, cancer incidence and mortality, and quality-adjusted life-years). Jurisdictions planning the implementation of population-based breast cancer screening can compare the impact of different screening strategies. For jurisdictions that have an organized breast cancer screening program in place, OncoSim-Breast could help investigate emerging issues such as increasing false-positives and customizing screening protocols based on different risk factors. In addition, jurisdictions can use the model to assess the impact of service disruptions during the COVID-19 pandemic. For example, they can estimate the impact of pausing screening for various time intervals on the stage of diagnosis and breast cancer deaths. They can also compare the impact of different strategies for restoring screening programs on downstream resources, such as follow-up diagnostics, biopsies and surgeries.

### Limitations

This paper has several limitations. First, OncoSim is a simulation model built using the best available data; the accuracy of projections depends on the quality of data input and the validity of assumptions. To address the issue of rapidly emerging evidence, OncoSim-Breast allows users to modify the inputs and assumptions. Second, our validation exercise comparing OncoSim-Breast’s projections with more recent Canadian Cancer Registry data was limited by the availability and quality of data in the Registry. Third, our simulation of the UK Age trial was an exploratory external validation exercise; we did not calibrate the model to match historically poorer breast cancer outcomes at that time. Fourth, OncoSim-Breast was built to be a multipurpose breast cancer simulation tool and could simulate many scenarios; therefore, it would not be feasible to validate all its possible projections against observed data. To ensure OncoSim-Breast’s relevance for supporting policy decisions, the team compares OncoSim-Breast’s projections with emerging real-world data and refines the model based on new evidence, on an ongoing basis. In the upcoming releases, examples of further enhancements will include adding emerging data on new screening modalities, costs of new treatments, and other factors that might affect screening performance, such as breast density and polygenic risk scores.

### Conclusion

OncoSim-Breast is a natural history-based simulation model developed using Canadian cancer incidence, mortality, screening program and cost data. It reproduces breast cancer trends in the Canadian Cancer Registry, breast cancer mortality in the Vital Statistics, and the breast cancer screening effects observed in a randomized screening trial.

## Data Availability

This paper describes a mathematical model that is available to public sector use at no charge. All model inputs are available in the model to authorized users.

http://www.oncosim.ca

## ACKNOWLEDGMENTS

OncoSim is led and supported by the Canadian Partnership Against Cancer, with model development by Statistics Canada, and is made possible through funding provided by Health Canada. We thank the many individuals who have contributed to the conceptualization, development, review, and application of OncoSim (see https://www.partnershipagainstcancer.ca/tools/oncosim/acknowledgements/). In particular, we would like to acknowledge the substantial contribution from other members of the OncoSim-Breast technical working group (full list is in Supplemental Appendix 8). Also, we would like to thank Dr. James Mainprize for his contribution in the Age trial validation exercise.

